# Real-Time Estimation of the Risk of Death from Novel Coronavirus (COVID-19) Infection: Inference Using Exported Cases

**DOI:** 10.1101/2020.01.29.20019547

**Authors:** Sung-mok Jung, Andrei R. Akhmetzhanov, Katsuma Hayashi, Natalie M. Linton, Yichi Yang, Baoyin Yuan, Tetsuro Kobayashi, Ryo Kinoshita, Hiroshi Nishiura

## Abstract

The exported cases of 2019 novel coronavirus (COVID-19) infection that were confirmed outside of China provide an opportunity to estimate the cumulative incidence and confirmed case fatality risk (cCFR) in mainland China. Knowledge of the cCFR is critical to characterize the severity and understand the pandemic potential of COVID-19 in the early stage of the epidemic. Using the exponential growth rate of the incidence, the present study statistically estimated the cCFR and the basic reproduction number—the average number of secondary cases generated by a single primary case in a naïve population. We modeled epidemic growth either from a single index case with illness onset on 8 December, 2019 (Scenario 1), or using the growth rate fitted along with the other parameters (Scenario 2) based on data from 20 exported cases reported by 24 January, 2020. The cumulative incidence in China by 24 January was estimated at 6924 cases (95% CI: 4885, 9211) and 19,289 cases (95% CI: 10,901, 30,158), respectively. The latest estimated values of the cCFR were 5.3% (95% CI: 3.5%, 7.5%) for Scenario 1 and 8.4% (95% CI: 5.3%, 12.3%) for Scenario 2. The basic reproduction number was estimated to be 2.1 (95% CI: 2.0, 2.2) and 3.2 (95% CI: 2.7, 3.7) for Scenarios 1 and 2, respectively. Based on these results, we argued that the current COVID-19 epidemic has a substantial potential for causing a pandemic. The proposed approach provides insights in early risk assessment using publicly available data.

## 1. Introduction

Since 8 December, 2019, clusters of pneumonia cases of unknown etiology have emerged in Wuhan City, Hubei Province, China [1,2]. Virological investigation suggests that the causative agent of this pneumonia is a novel coronavirus (COVID-19) [3]. As of 27 January, 2020, a total of 4515 cases including 106 deaths were confirmed [4]. Forty-one cases of COVID-19 infections were also reported outside China, in other Asian countries, the United States, France, Australia, and Canada.

A local market selling seafood and wildlife in Wuhan was visited by many cases in the initial cluster, indicating that a common-source zoonotic exposure may have been the main mode of transmission [5]. However, even after shutting down the market, the number of cases continued to grow across China and several instances of household transmission were reported [6]. It is now speculated that sustained human-to-human transmission aided in the establishment of the epidemic [7] and that reported case counts greatly underestimated the actual number of infections in China [8].

Early assessment of the severity of infection and transmissibility can help quantify the pandemic potential of COVID-19 and anticipate the likely number of deaths by the end of the epidemic. One important epidemiological measure of severity is case fatality risk (CFR), which can be measured with three different approaches, by estimating (i) the proportion of the cumulative number of deaths out of the cumulative number of cases at a point in time, (ii) the ratio of the cumulative number of deaths to the cumulative number of infected individuals whose clinical outcome is known (i.e., the deceased or recovered), and (iii) the risk of death among confirmed cases, explicitly accounting for the time from illness onset to death [9]. Estimating the CFR using the ratio of deaths to confirmed cases (cCFR), with an adjustment of the time delay from illness onset to death (i.e., method (iii)), can provide insight into severity of the disease, because the naïve CFR based on method (i) tends to be underestimated due to the real-time nature of the growth of fatal cases. For example, during the early stage of an epidemic, failing to right-censor cases with respect to the time delay from illness onset to death may lead to underestimation of the CFR. This is because death due to infection may yet occur following case identification [9–11].

When the CFR denominator only includes confirmed cases it is referred to as the cCFR [10] and may overestimate the actual CFR among all infected individuals due to under-ascertainment of infections in the population. Nonetheless, the cCFR is still a valuable measure of the upper bound of the symptomatic CFR (sCFR) among all symptomatic cases, particularly in circumstances of high uncertainty, such as the emergence of a new human pathogen (i.e., COVID-19).

The basic reproduction number (*R*_0_), the average number of secondary cases generated by a single primary case in a fully susceptible population, represents an epidemiological measurement of the transmissibility, helping us to quantify the pandemic potential of COVID-19. Here, we define a pandemic as the worldwide spread of a newly emerged disease, in which the number of simultaneously infected individuals exceeds the capacity for treatment [12]. Using the growth rate of the estimated cumulative incidence from exportation cases and accounting for the time delay from illness onset to death, the present study aims to estimate the cCFR of COVID-19 in real-time.

## 2. Methods

## 2.1. Epidemiological Data

Information on exported COVID-19 cases who were confirmed in other countries and deaths due to COVID-19 infection in China were retrieved from the first announcement date of the current outbreak (i.e., 31 December, 2019) through 24 January, 2020. The cutoff time of 24 January, 2020 was specifically selected to reflect the governmental ban on public transportation from Wuhan (including flights) which was invoked on 23 January, 2020 [13]. All data were collected either from government websites or from media quoting government announcements. Our data include 51 cases diagnosed outside China who had illness onset through 24 January, 2020 and were reported by 9 February, 2020 and the dates of illness onset and death among 41 deceased cases in China.

### 2.2. Estimation of the Delay Distributions

The observed incidence *i*(*t*) by date of illness onset *t* is modeled using an exponential growth model with the rate *r*: *i*(*t*) = *i*_0_*e*^*rt*^, where *i*_0_ is the expected number of infected cases at time *t* = 0. The cumulative incidence *I*(t) is an integral of *i*(*t*) over the time interval from zero to *t* that can be written as 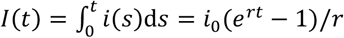. The cumulative incidence is adjusted to the date of report by the factor *u* dependent on the parameters of the delay distribution. For the estimation of time delay distribution from illness onset to death, we accounted for right truncation and modeled and used a lognormal distribution with parameters adopted from an earlier study [14]. Let *f*(*t*; *θ*) be the lognormal distribution with parameters *θ*_*d*_ = {*a*_*d*_, *b*_*d*_}. Then, the cumulative incidence *I(t)* by date of report *t* can be adjusted to the time from illness onset to death and report by simply multiplying it by the factor *u*(*r, θ*_*d*_), which is a consequence of the exponentially growing epidemic [10]. The factor *u* is defined by the following integral:

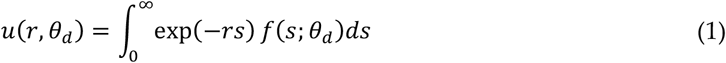

The adjusted cumulative incidence then obeys *u*(*r, θ*_*d*_). *I*(*t*). The cumulative number of deaths *D*(*t*) reported by date *t* is the result of Binomial sampling:

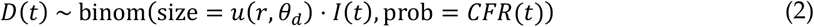

Let *a*_*e*_ and *b*_*e*_ be the shape and inverse scale of the gamma distribution modeling the distribution of time interval from illness onset to the report for observed exported cases. The cumulative incidence in China can be then adjusted by the multiplicative factor ũ(*r, θ*_*e*_ = {*a* _*e*_, *b* _*e*_}). The number of exported cases *E*(*t*) can then *be sampled from another Binomial distribution*:

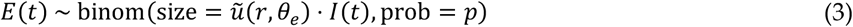

where *p* describes the probability of finding a traveler from Wuhan among all travelers from China subject to the detection time window of the virus *T* = 12.5 days [8]. Given that the total volume of inbound passengers from China is *M* = 5.56 million passengers per year, the fraction of Wuhan travelers is *ϕ* = 0.021%, and the population of Wuhan is *n* = 11 million, the probability *p* is given by

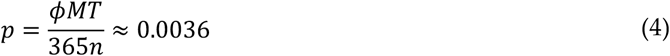

### 2.3. Statistical Inference

First, we fitted the delay distribution of the time from illness onset to report Δ*T*_*e,k*_ (*k* ≤ *K*_*e*_) to the gamma distribution. We define the log-likelihood as follows:

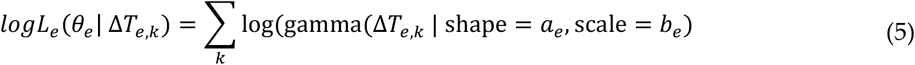

that is maximized to find the mean values of the parameters 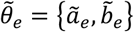, used in the following step.

Second, we fitted the observed counts of exported cases and deaths by considering two other likelihoods respectively to each process:

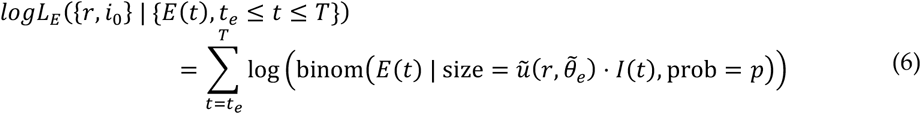

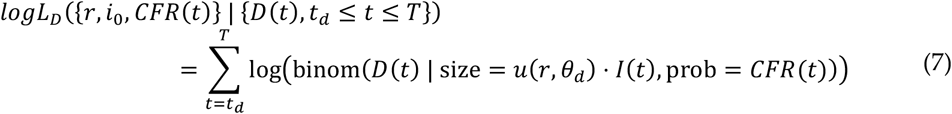

where *t*_*e*_ and *t*_*d*_ are the times at which the first exportation event and the first death are observed. The total log-likelihood is given by the sum of two log-likelihoods:

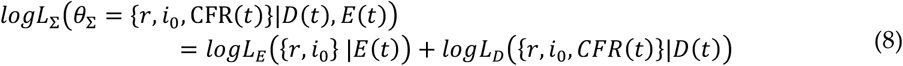

which is then maximized to determine the best-fit parameters {*r, i*_0_, *CFR*(*t*)}.

We considered two possible scenarios to fit to the data. In the first scenario (refers to “Scenario 1”), the parameter *i*_0_ is fixed at one on the date of illness onset for the first COVID-19 confirmed case (i.e., 8 December, 2019), providing a fixed starting point for the exponential growth of the cumulative incidence. Although 8 December, 2019 is assumed as the starting point for exponential growth of the cumulative incidence in Scenario 1, there is still uncertainty using this date as a proxy for the beginning of exponential growth due to inconsistencies reported on illness onset for the earliest reported cases [2,5]. Therefore, we conducted a sensitivity analysis where the start date for exponential growth was varied between 1 December and 10 December, 2019. In the second scenario (refers to “Scenario 2”), all parameters {*r, i*_0_, *CFR*(*t*)} are variable, and calculation begins on the date the first exported case was observed (i.e., 13 January, 2020). For both scenarios, we conducted sensitivity analyses that estimated the growth rate, cumulative incidence, and cCFR value using different cut-off times, values of the detection window *T*, and sizes of the catchment population of the Wuhan International airport *n*.

The value of the basic reproduction number *R*_0_ for the COVID-19 epidemic was calculated as

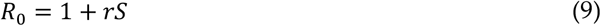

where *r* is the estimated growth rate for each scenario and *S* is the mean serial interval for successive COVID-19 infections. The value of the serial interval was adopted from a published study [2], and a gamma distribution was fitted to six known pairs of the infectee and infector (mean = 7.5 days, standard deviation (SD) = 3.4 days).

In our analysis, we employed Markov chain Monte Carlo (MCMC) simulations with eight chains of 4250 samples each, with 2500 samples used for the tuning stage, leveraging the No-U-Turn sampler (NUTS) part of the Python PyMC3 package. However, one of the input variables for the Binomial distribution ũ(*r, θ*_*e*_)· *I*(*t*) or *u*(*r, θ*_*d*_). *I*(*t*) was modeled with a continuous variable in our approach. We used the gamma distribution, matching the first two moments to obtain the transformation of the discrete Binomial distribution to its continuous approximation [16]. We avoided joint estimation of all parameters {*θ*_Σ_, *θ*_*e*_, *θ*_*d*_} due to heterogeneity in the aggregated data—a similar issue is discussed in [10]. Instead, we implemented a sequential fitting: first we considered only the likelihood *L*_*e*_, then we fit the likelihood *L*_Σ_ using the mean values of the estimated parameters obtained in the previous round. Finally, we verified the obtained fit by calculating pointwise estimates using maximum likelihood estimation with confidence intervals derived as profile likelihood-based intervals. We found that both approaches were at complete agreement in their results.

## 3. Results

Figures 1A,B show the mean and SD of the time from illness onset to reporting and death, respectively. Employing the gamma distribution, the mean time from illness onset to reporting was estimated at 7.1 days (95% CI: 5.9, 8.4). The mean time from illness onset to death—adopted from a previous study [14]—was estimated at 20.2 days (95% CI: 15.1, 29.5), which led to the lognormal distribution with location parameter of 2.84 and scale parameter of 0.52.

**Figure 1.**
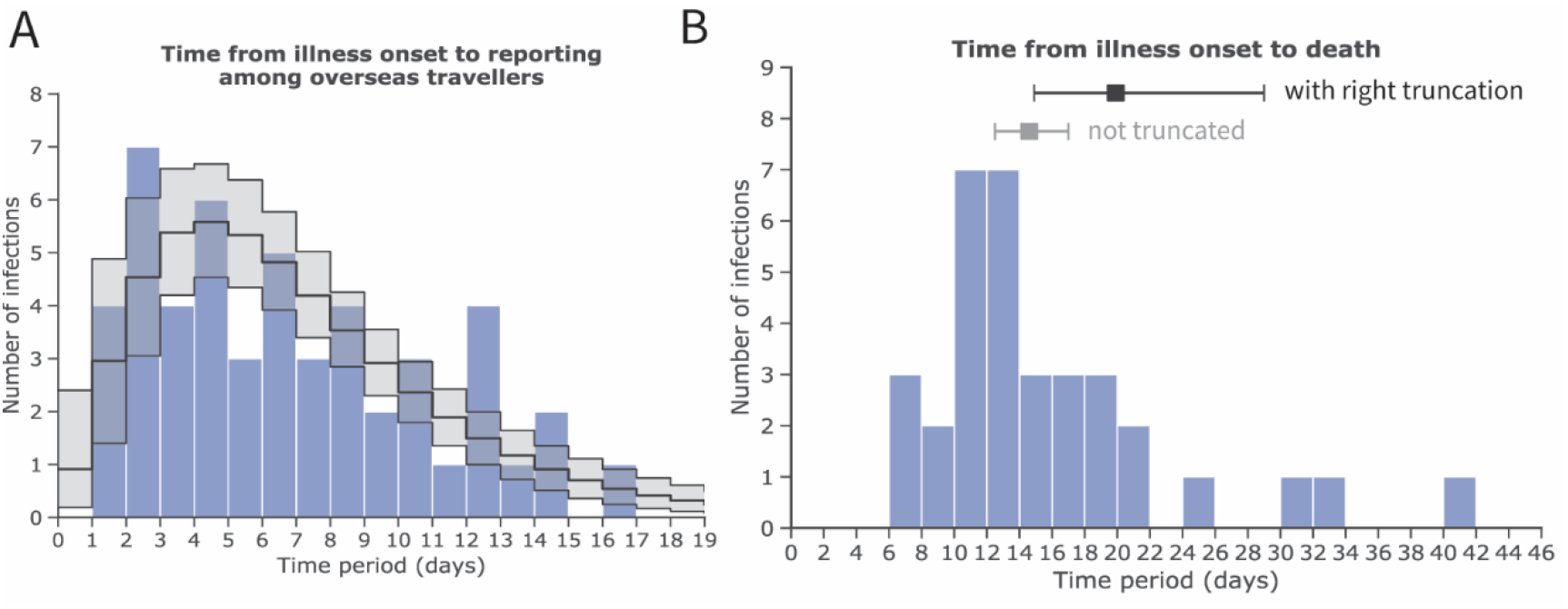
Estimates of the mean and standard deviation (SD) of the time from illness onset to reporting and death cases, accounting for right truncation, with novel coronavirus (COVID-19) infection in China, 2020. Inference of A and B was conducted among (**A**) exported cases observed in other countries and (**B**) deceased cases in China. (**A**) Frequency distribution of the time from illness onset to reporting among exported cases employing a gamma distribution with a mean of 7.1 days (95% CI: 5.9, 8.4) and SD of 4.4 days (95% CI: 3.5, 5.7). (**B**) Frequency distribution of the time from illness onset to death with a mean of 19.9 days (95% CI: 14.9, 29.0) (shown in black) and SD of 11.4 days (95% CI: 6.5, 21.6) employing a lognormal distribution and accounting for right truncation. For reference, the estimate of the mean and its 95% credible intervals without accounting for right truncation are shown in grey. The values for distribution of time from illness onset to death are adopted from an earlier study [14]. The blue bars show empirically observed data collected from governmental reports (as of 24 January, 2020).

Subsequently, the cumulative incidence was estimated from exported case data by fitting an exponentially growing incidence curve for both Scenarios 1 and 2 (Figure 2). As of 24 January, 2020, 20 exported cases were reported, and the cumulative incidence in China was estimated at 6924 cases (95% CI: 4885, 9211) in Scenario 1 and 19,289 cases (95% CI: 10,901, 30,158) in Scenario 2. Table 1 shows the real-time update of the estimated cumulative incidence. The exponential growth rates (*r*), derived from the growth rate of cumulative incidence, were estimated at 0.15 per day (95% CI: 0.14, 0.15) and 0.29 per day (95% CI: 0.22, 0.36) in Scenarios 1 and 2, respectively.

**Table 1.** Exportation events and estimated incidence in China, 2020.

| Importing Locations | Date of Report | Cumulative Count | Estimated Incidence in China (95% CI) |  |
| --- | --- | --- | --- | --- |
|  |  |  | Scenario 1 | Scenario 2 |
| Thailand | 13 January 2020 | 1 | 1828 (1397, 2288) | 1369 (1003, 1782) |
| Japan | 16 January 2020 | 2 | 2120 (1605, 2672) | 1829 (1392, 2309) |
| Thailand | 17 January 2020 | 3 | 2458 (1845, 3119) | 2444 (1894, 3033) |
| South Korea | 20 January 2020 | 4 | 3832 (2802, 4962) | 5882 (4252, 7629) |
| Taiwan, United States | 21 January 2020 | 6 | 4443 (3220, 5792) | 7901 (5425, 10,662) |
| Thailand | 22 January 2020 | 8 | 5151 (3700, 6761) | 10,626 (6897, 15,003) |
| Singapore, Vietnam | 23 January 2020 | 11 | 5972 (4252, 7892) | 14,308 (8661, 21,250) |
| Japan, Nepal, South Korea, Singapore, Thailand, United States | 24 January 2020 | 20 | 6924 (4885, 9211) | 19,289 (10,901, 30,158) |

CI, confidence interval (the 95% CI was derived from the Markov chain Monte Carlo method).
Scenario 1 indicates the estimated exponential growth rate with the assumed illness onset date of the first COVID-19 case (i.e., 8 December, 2019), while Scenario 2 presents the estimated exponential growth rate from the date of first exportation event (i.e., 13 January, 2020).

**Figure 2.**
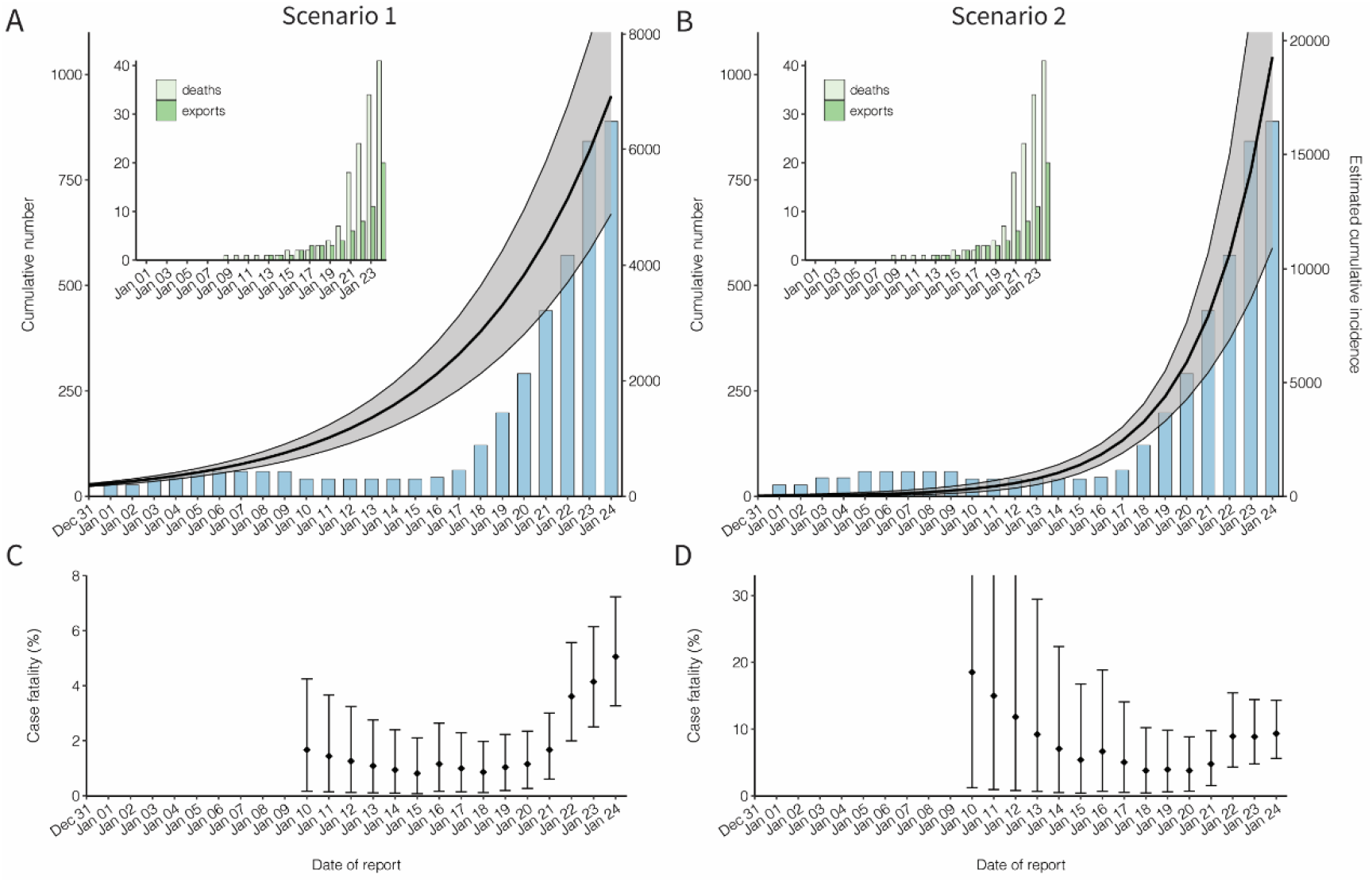
Cumulative incidence and the confirmed case fatality risk of the novel coronavirus (COVID-19) outbreak in China, 2020. (**A**,**B**) Observed and estimated cumulative number of cases in China by the date of report. An exponential growth curve was extrapolated using the exported case data. Scenario 1 extrapolated the exponential growth from December to first case on 8 December, 2019, while Scenario 2 started the estimation of the exponential growth only from 13 January, 2020. The black line and shaded area represent median and 95% credible interval of the cumulative incidence in China, respectively. The blue bars show the cumulative number of reported cases from the government of mainland China. The cumulative number of reported cases was not used for fitting, but it was shown for comparison between the cumulative number of reported and estimated cases in China. There is a decrease in the cumulative number of reported cases in early January, because only 41 cases tested positive for the novel coronavirus among the reported 59 cases on 10 Jan, 2020. Left-top panels on both **A** and **B** show the cumulative numbers of exported cases observed in other countries and the cumulative number of deaths in China, represented by dark and light green bars, respectively. (**C**,**D**) Confirmed case fatality risk (cCFR) by the date of reporting. Each value of cCFR was estimated as the ratio of cumulative number of estimated incidence to death at time *t*. The points and error bars represent median and its 95% credible interval of cCFR. All 95% credible intervals were derived from Markov chain Monte Carlo simulations.

CI, confidence interval (the 95% CI was derived from the Markov chain Monte Carlo method). Scenario 1 indicates the estimated exponential growth rate with the assumed illness onset date of the first COVID-19 case (i.e., 8 December, 2019), while Scenario 2 presents the estimated exponential growth rate from the date of first exportation event (i.e., 13 January, 2020).

Figures 2C and 2D show the estimated cCFR value accounting for the time delay from illness onset to death under Scenarios 1 and 2, respectively. A total of 41 confirmed deaths were reported as of 24 January, 2020, the cCFR value was estimated at 5.3% (95% CI: 3.5%–7.5%) for Scenario 1 and 8.4% (95% CI: 5.3%–12.3%) for Scenario 2, respectively.

We estimated the basic reproduction number for the COVID-19 infection, using the estimated exponential growth (*r*) and accounting for possible variations of the mean serial interval (Figure 3). Assuming that the mean serial interval was 7.5 days [2], the basic reproduction number was estimated at 2.10 (95% CI: 2.04, 2.16) and 3.19 (95% CI: 2.66, 3.69) for Scenarios 1 and 2, respectively. However, as the mean serial interval varies, the estimates can range from 1.6 to 2.6 and 2.2 to 4.2 for Scenarios 1 and 2, respectively.

**Figure 3.**
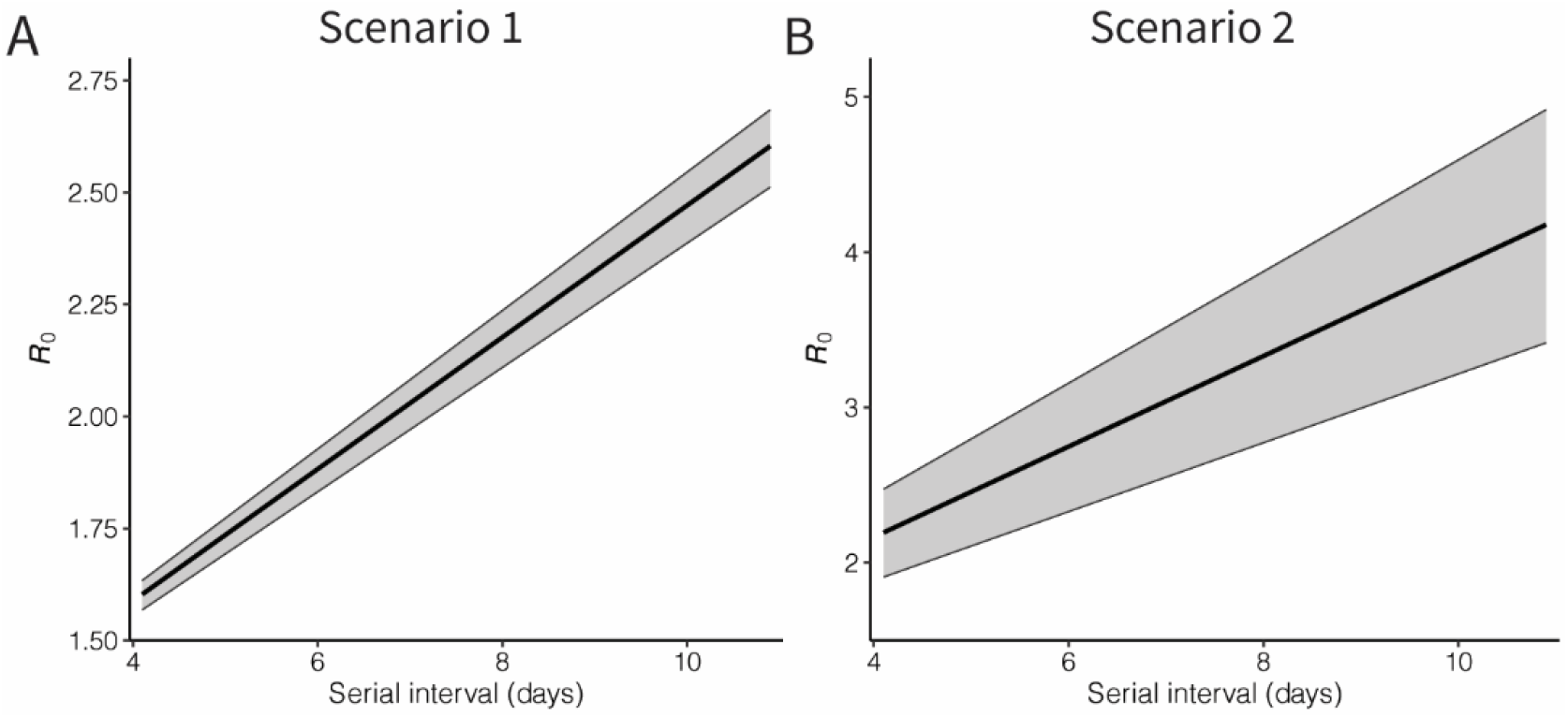
Basic reproduction number of novel coronavirus (COVID-19) infections in China, 2020. Black lines and grey shades represent the median and 95% credible intervals of the basic reproduction number. Panel **A** shows the result of Scenario 1, in which an exponential growth started from the assumed illness onset date of index case, while Panel **B** shows the result from exponential growth from the first exported case (Scenario 2). The 95% credible intervals were derived from Markov chain Monte Carlo method.

To address the uncertainty in the unobserved date of illness onset of the index case in Scenario 1, cCFR was estimated by varying the starting date of the exponential growth in the incidence by placing the single index case between 1 and 10 December, 2019 (Figure S1 and Table S1). When we assumed the date of illness onset of the index case was 1 December, 2019, the estimated incidence in China and the cCFR on 24 January, 2020 were estimated at 4718 (95% CI: 3328, 6278) and 5.3% (95% CI: 3.5, 7.6). The sensitivity analyses for varied cutoff dates between 15 and 24 January, 2020 were conducted. Depending on the number of time points, the estimates of the cumulative incidence and cCFR have similar values in Scenario 1 (Figure S2) but slightly decreased in Scenario 2, when the cutoff date was earlier (Figure S3). In addition, considering the uncertainty of two fixed parameters (i.e., detection time window and catchment population in Wuhan airport), sensitivity was assessed by varying those parameters. As the catchment population increases and the detection window time decreases, estimated cCFR on 24 January, 2020 gets smaller due to increased number of incidence in China (Table S2 and S3).

## 4. Discussion

The present study estimated the risk of death among confirmed cases while addressing ascertainment bias by using data from cases diagnosed outside mainland China and a right-censored likelihood for modeling the count of deceased cases. The estimated cCFR value was 5.3% [3.5, 7.5] when the date of illness onset for the index case was fixed a priori at 8 December, 2019 (Scenario 1), and 8.4% [5.3, 12.3%] when the timing of the exponential growth of epidemic was fitted to data alongside with other model parameters (Scenario 2). The estimated value of *u* in Scenario 2, which adjusts the time delay from illness onset to death, was smaller than that in Scenario 1 due to the larger value of the growth rate of incidence, while the estimated cCFR value as of 24 January, 2020 was larger in Scenario 2 compared with Scenario 1. Depending on the available data, the estimate of the cCFR (i.e., the delay-adjusted risk of death among confirmed cases) may vary in time and show some fluctuations. In addition to that, we estimated the value of the basic reproduction number *R*_0_ in the range of 1.6–2.6 for Scenario 1 and 2.2–4.2 for Scenario 2. From either estimate, we conclude that COVID-19 has substantial potential to spread via human-to-human transmission. However, *R*_0_ > 1 does not guarantee that a single exported (and untraced) case would immediately lead to a major epidemic in the destination country as government responses such as border control, isolation of suspected cases, and intensive surveillance should serve to reduce opportunities for transmission to occur [17–19].

Our cCFR estimates of 5.3% and 8.4% indicate that the severity of COVID-19 is not as high as that of other diseases caused by coronaviruses, including severe acute respiratory syndrome (SARS), which had an estimated CFR of 17% in Hong Kong [9,10,20], and Middle East respiratory syndrome, which had an estimated CFR of 20% in South Korea [21]. Nonetheless, considering the overall magnitude of the ongoing epidemic, a 5%–8% risk of death is by no means insignificant. In addition to quantifying the overall risk of death, future research must identify groups at risk of death (e.g., the elderly and people with underlying comorbidities) [22,23]. Moreover, considering that about 9% of all infected individuals are ascertained and reported [24], the infection fatality risk (IFR), i.e., the risk of death among all infected individuals, would be on the order of 0.5% to 0.8%.

Our estimated *R*_0_ range of 1.6–4.2 for COVID-19 is consistent with other preliminary estimates posted on public domains [25–28], and is comparable to the *R*_0_ of SARS, which was in the range of 2–5 during the 2003 outbreak in Singapore [15]. Between our two estimates, the latter scenario yielded a greater value than the former, and there was an increasingly improved ascertainment in early January 2020. The virus was identified and sequenced on 7 January, 2020 and subsequently, the primer was widely distributed, allowing for rapid laboratory identification of cases and contributing to a time-dependent increase in the number of confirmed cases out of China. Consequently, Scenario 2, which was fully dependent on the growth rate of exported cases, could have overestimated the intrinsic growth rate of cases. Considering the estimated value of *R*_0_, the possibility of presymptomatic transmission in the ongoing epidemic is a critical question, as it would have a substantial impact on public health response to the epidemic (e.g., whether the contact tracing should be prioritized or not) as well as overall predictability of the epidemic during the containment stage [29,30].

From the technical side, it should be emphasized that our proposed approach can be especially useful during the early stage of an epidemic when local surveillance is affected by substantial ascertainment bias and export and death data are available and better ascertained. Nonetheless, caution must be used when implementing similar estimations for the COVID-19 epidemic, as all flights from Wuhan airport were grounded as of 23 January, 2020 [13] and this intervention abruptly changed the human migration network. Despite the decrease in the outbound flow of travelers from Wuhan, there is a substantial risk that the next epidemic wave will originate from other cities.

There are five main limitations in the present study. First, our results present an estimate for the cCFR which only addresses fatality among confirmed cases. More precise IFR estimate that includes infected individuals other than confirmed cases can only be estimated using additional pieces of data (e.g., seroepidemiological data or outpatient clinic visits). It should be noted that not only the denominator but also numerator values are also subject for better estimation (e.g. excess mortality estimate). Second, our study relied on limited empirical data that were extracted from publicly available data sources. Thus, future studies with greater sample size and precision are needed. Nonetheless, we believe that this study will improve the situational assessment of the ongoing epidemic. Third, our assumed date of illness onset for the index case in Scenario 1 is based on initial reports of the earliest onset date for a case, and the continued exponential growth with the rate *r* is the authors’ extrapolation. However, we conducted a sensitivity analysis and ensured that the resulting statistical estimates would not greatly vary from our main results. Fourth, there is an uncertainty in the detection window time *T*. Since the epidemiological investigations are being actively implemented outside China, we believe that the sum of the incubation period and the infectious period can be a plausible estimation of the detection time window. Fifth, heterogeneous aspects of death (e.g. age and risk groups) need to be addressed in the future studies.

In conclusion, the present study has estimated cCFR to be on the order of 5%–8% and *R0* to be 1.6–4.2, endorsing the notion that COVID-19 infection in the ongoing epidemic possesses the potential to become a pandemic. The proposed approach can also help direct risk assessment in other settings with the use of publicly available datasets.

## Data Availability

All data were collected either from government websites or media quoting government announcements.

## Supplementary Materials

The following are available online at www.mdpi.com/xxx/s1, Figure S1: Sensitivity analysis with varying the start point of exponential growth in cumulative incidence from 1 to 10 December, 2019, Figure S2: Sensitivity analysis with varying the cutoff date from 15 to 24 January, 2020 in estimation Scenario 1, Figure S3: Sensitivity analysis with varying the cutoff date from 15 to 24 January, 2020 in estimation Scenario 2, Table S1: Sensitivity analysis with varying the start point of exponential growth in cumulative incidence from 1 to 10 December, 2019 in estimation Scenario 1, Table S2: Sensitivity analysis with varying the catchment population size in Wuhan airport, Table S3: Sensitivity analysis with varying the detection time window.

### Author contributions

S.-m.J., A.R.A., and H.N. conceived the study and participated in the study design. All authors assisted in collecting the data. S.-m.J. and A.R.A. analyzed the data and S.-m.J., A.R.A., K.H. and H.N. drafted the manuscript. All authors edited the manuscript and approved the final version.

### Funding

H.N. received funding from the Japan Agency for Medical Research and Development (AMED) [grant number: JP18fk0108050]; the Japan Society for the Promotion of Science (JSPS) KAKENHI [grant numbers, H.N.: 17H04701, 17H05808, 18H04895 and 19H01074; R.K.: 18J21587], the Inamori Foundation, and the Japan Science and Technology Agency (JST) CREST program [grant number: JPMJCR1413]. S.M.J. and N.M.L. receive graduate study scholarships from the Ministry of Education, Culture, Sports, Science and Technology, Japan. B.Y. wishes to thank China Scholarship Council.

## Supplementary materials

### A. Sensitivity analysis by varying the starting point of exponential growth of cumulative novel coronavirus incidence from 1 December to 10 December, 2019

The starting point of exponential growth of cumulative case incidence was fixed as the illness onset date of the index case (i.e, 8 December 2019) in Scenario 1, while it was allowed to vary from the day the first exportation case was observed (i.e., 13 January 2020) in Scenario 2. However, since there is uncertainty regarding the actual starting point of exponential growth in Scenario 1 due to discrepancies in the date of illness onset for the first reported case, a sensitivity analysis was conducted by varying this date from 1 December 2019 to 10 December 2019. Table S1 and Figure S1 show the estimated growth rate, cumulative incidence, and cCFR that result from this analysis.

**Table S1.** Sensitivity analysis with varying the start point of exponential growth in cumulative incidence from 1**–**10 December, 2019 in estimation Scenario 1.

| The start point of exponential growth | Exponential growth rate (95% CI) | Estimated incidence in China on 24 January (95% CI) | cCFR on 24 January (95% CI) |
| --- | --- | --- | --- |
| 1 December | 0.12 (0.11, 0.12) | 4718 (3328, 6278) | 5.33% (3.50, 7.58) |
| 2 December | 0.12 (0.11, 0.13) | 4896 (3473, 6472) | 5.35% (3.57, 7.61) |
| 3 December | 0.12 (0.12, 0.13) | 5087 (3584, 6733) | 5.40% (3.59, 7.62) |
| 4 December | 0.13 (0.12, 0.13) | 5392 (3730, 7024) | 5.44% (3.62, 7.68) |
| 5 December | 0.13 (0.12, 0.14) | 5506 (3908, 7309) | 5.48% (3.66, 7.74) |
| 6 December | 0.14 (0.13, 0.14) | 5733 (4067, 7583) | 5.53% (3.68, 7.80) |
| 7 December | 0.14 (0.13, 0.15) | 6089 (4317, 8099) | 5.52% (3.68, 7.76) |
| 8 December | 0.14 (0.14, 0.15) | 6266 (4435, 8285) | 5.64% (3.74, 7.96) |
| 9 December | 0.15 (0.14, 0.16) | 6565 (4663, 8687) | 5.69% (3.79, 8.03) |
| 10 December | 0.15 (0.14, 0.15) | 6924 (4885, 9211) | 5.27% (3.51, 7.45) |

cCFR, confirmed case fatality risk; CI, confidence interval (the 95% CI was derived from the Markov chain Monte Carlo method).

**Figure S1.**
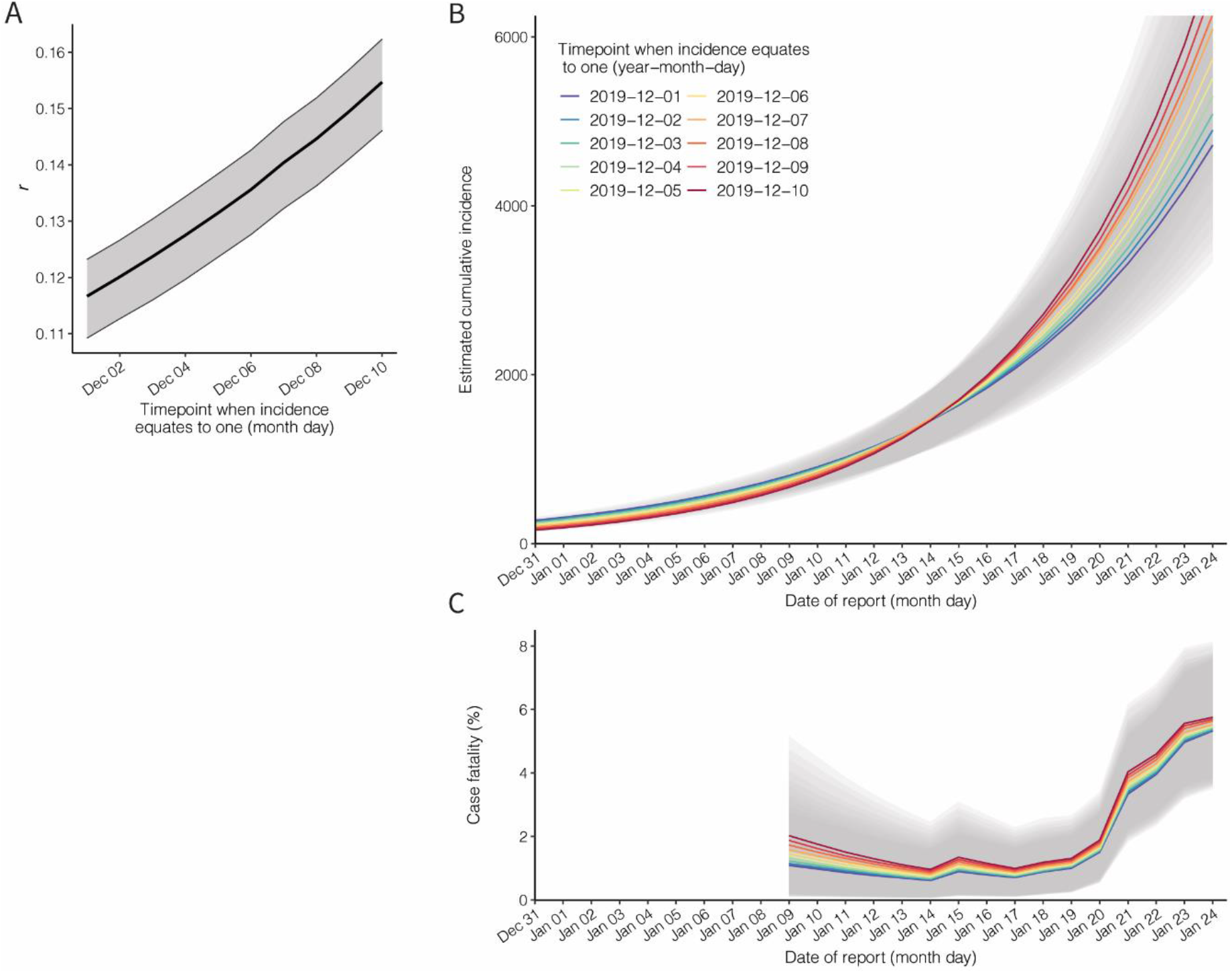
Sensitivity analysis with varied starting point for exponential growth of cumulative case incidence from 1–10 December, 2019. The estimated values of the: A) exponential growth rate, B) cumulative incidence and C) confirmed case fatality risk are shown by varying the start date of exponential growth in Scenario 1. Each line and shaded area present the estimate and its 95% confidence interval (derived from the Markov chain Monte Carlo method).

### B. Sensitivity analyses by varying the data cutoff date from 15 January to 24 January, 2020

As the present study relies on real-time data, the estimates (i.e., growth rate, cumulative incidence, and cCFR) using different number data cutoff dates are shown in Figure S2 (Scenario 1) and S3 (Scenario 2).

**Figure S2.**
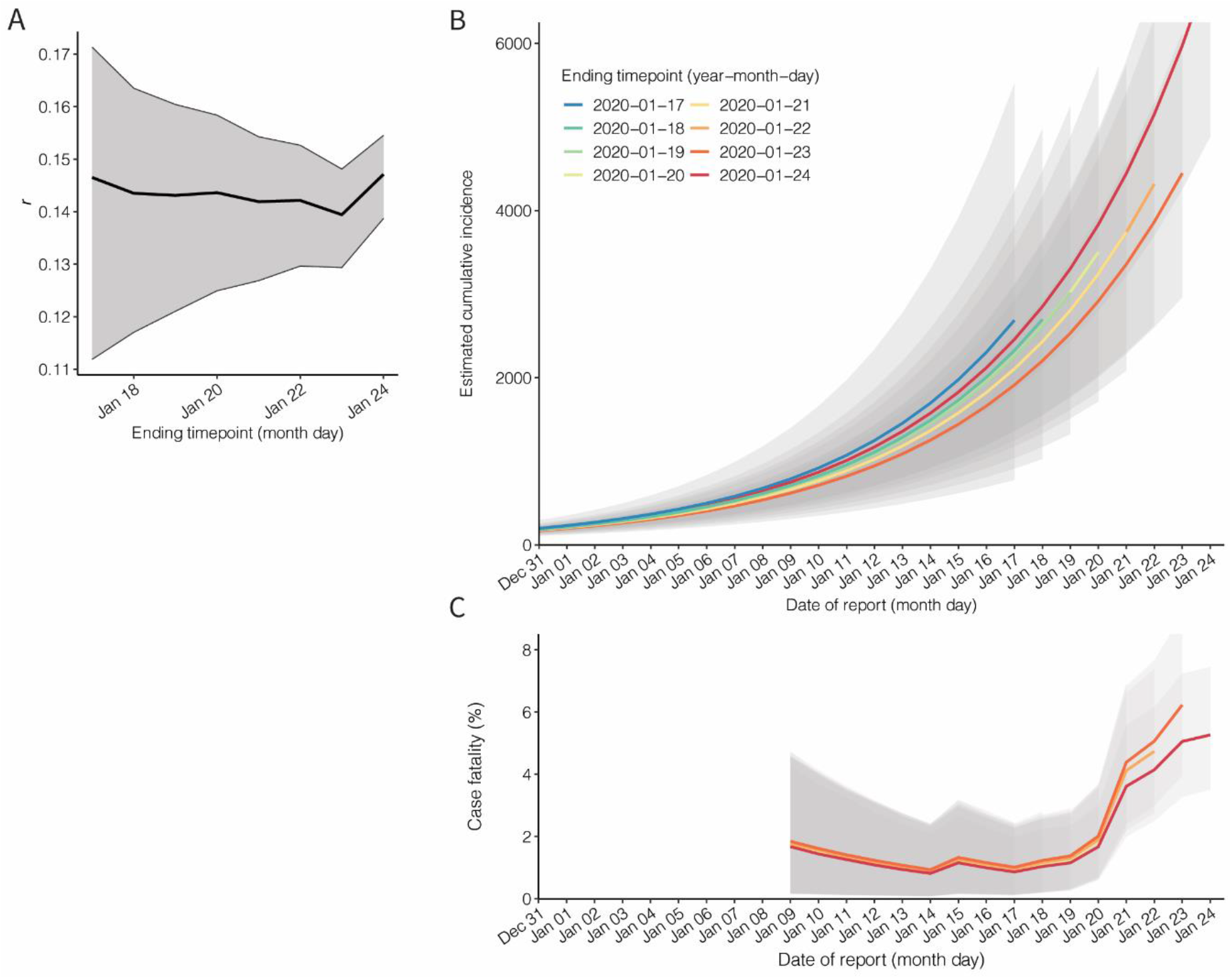
Sensitivity analysis in Scenario 1, varying the data cutoff date between 15 and 24 January 2020. The estimated values of the A) exponential growth rate, B) cumulative incidence, and C) confirmed case fatality risk depend on the data cutoff date (i.e., end point of each estimation). Each line and shaded area indicate the estimate and its 95% confidence interval.

**Figure S3.**
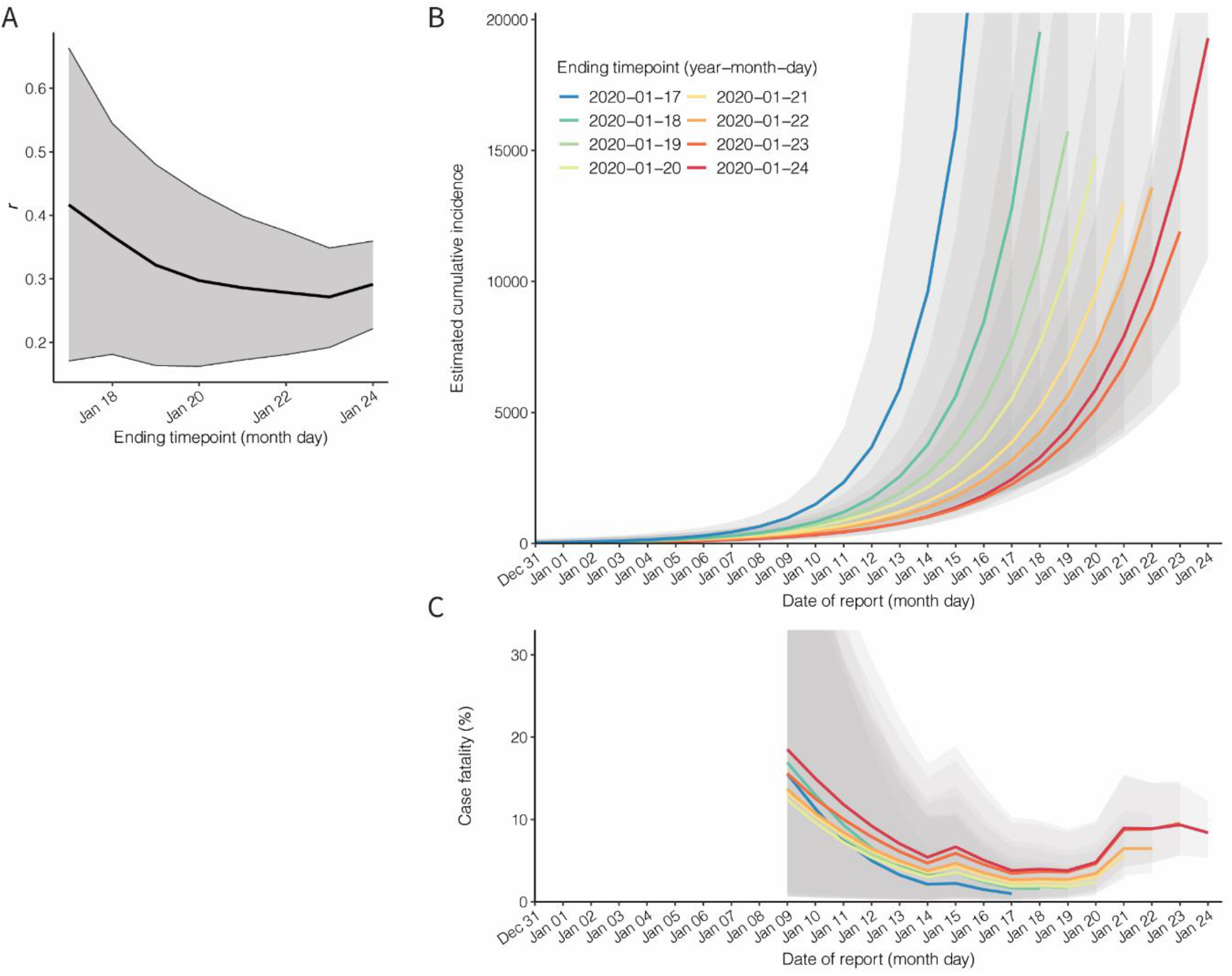
Sensitivity analysis in Scenario 2, varying the data cutoff date between 15 and 24 January 2020. The estimated values of the A) exponential growth rate, B) cumulative incidence, andC) confirmed case fatality risk depend on the data cutoff date (i.e., end point of each estimation). Each line and shaded area indicate the estimate and its 95% confidence interval.

### C. Sensitivity analyses by varying the detection time window (*T*) and catchment population size in Wuhan airport (*n*)

In the present study, the detection time window was fixed at 12.5 days, based assumed values for the incubation and infectious periods. The catchment population in Wuhan airport was also fixed at 11 million. As part of the sensitivity analysis, Table S2 shows the estimated growth rate, cumulative incidence and cCFR by varying only detection time window, while Table S3 shows the estimates with different catchment population sizes and a fixed detection time window for Scenarios 1 and 2, respectively.

**Table S2.** Sensitivity analysis varying the detection time window (*T*) with a catchment population in Wuhan airport of 11 million

| Scenario | Detection window time | Exponential growth rate (95% CI) | Estimated incidence on 24 January (95% CI) | cCFR on 24 January (95% CI) |
| --- | --- | --- | --- | --- |
| 1 | 3.6 days | 0.18 (0.18, 0.19) | 30094(21524, 39513) | 1.82% (1.20, 2.58) |
| 1 | 7.5 days | 0.16 (0.15, 0.17) | 12024 (8642, 15838) | 3.54% (2.36, 4.99) |
| 1 | 10 days | 0.15 (0.14, 0.16) | 8343 (5920, 11040) | 4.61% (3.06, 6.48) |
| 1 | 12.5 days | 0.15 (0.14, 0.15) | 6924 (4885, 9211) | 5.27% (3.51, 7.45) |
| 2 | 3.6 days | 0.33 (0.25, 0.40) | 80925 (45517, 127347) | 2.77% (1.72, 4.14) |
| 2 | 7.5 days | 0.31 (0.23, 0.38) | 34795 (19653, 54114) | 5.31% (3.37, 7.87) |
| 2 | 10 days | 0.30 (0.23, 0.37) | 26042 (14602, 41034) | 6.64% (4.24, 9.82) |
| 2 | 12.5 days | 0.29 (0.22, 0.36) | 19289 (10901, 30158) | 8.39% (5.34, 12.26) |

cCFR, confirmed case fatality risk; CI, confidence interval (the 95% CI was derived from Monte Carlo Markov Chain method).

**Table S3.**
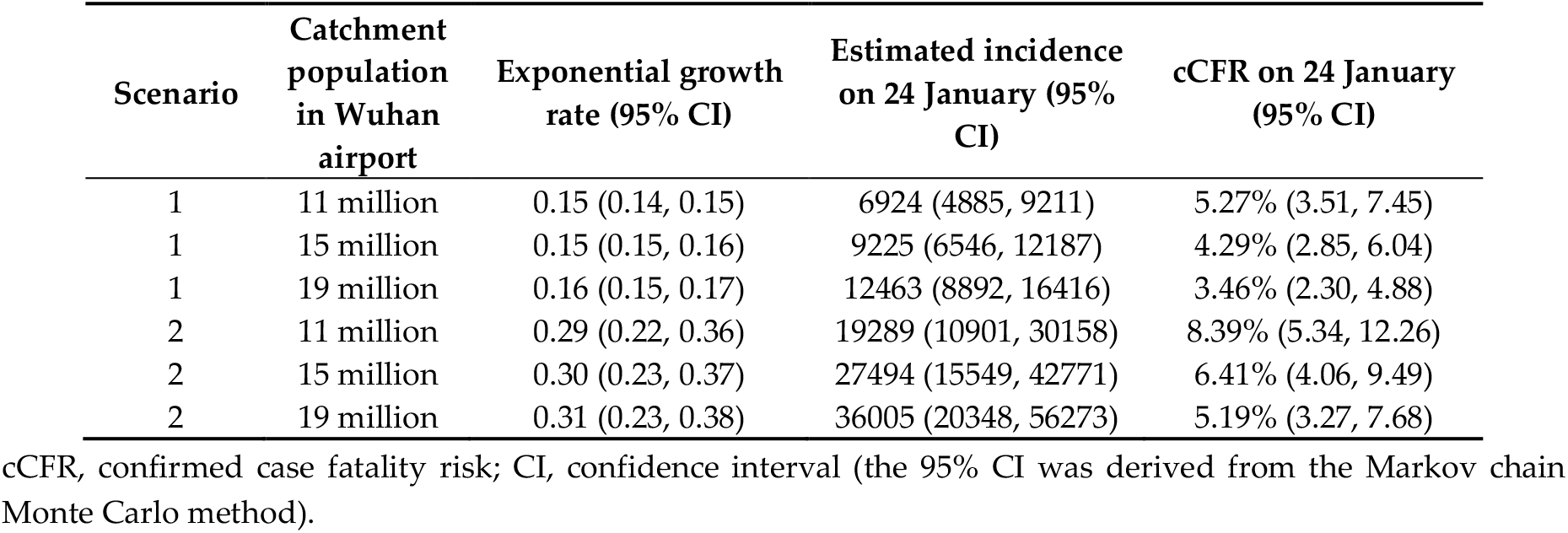
Sensitivity analysis by varying the catchment population size in Wuhan airport (*n*) with a fixed detection time window of 12.5 days

| Scenario | Catchment population in Wuhan airport | Exponential growth rate (95% CI) | Estimated incidence on 24 January (95% CI) | cCFR on 24 January (95% CI) |
| --- | --- | --- | --- | --- |
| 1 | 11 million | 0.15 (0.14, 0.15) | 6924 (4885, 9211) | 5.27% (3.51, 7.45) |
| 1 | 15 million | 0.15 (0.15, 0.16) | 9225 (6546, 12187) | 4.29% (2.85, 6.04) |
| 1 | 19 million | 0.16 (0.15, 0.17) | 12463 (8892, 16416) | 3.46% (2.30, 4.88) |
| 2 | 11 million | 0.29 (0.22, 0.36) | 19289 (10901, 30158) | 8.39% (5.34, 12.26) |
| 2 | 15 million | 0.30 (0.23, 0.37) | 27494 (15549, 42771) | 6.41% (4.06, 9.49) |
| 2 | 19 million | 0.31 (0.23, 0.38) | 36005 (20348, 56273) | 5.19% (3.27, 7.68) |

cCFR, confirmed case fatality risk; CI, confidence interval (the 95% CI was derived from the Markov chain Monte Carlo method).

